# Evaluation of Remote Monitoring Technology across different skin tone participants

**DOI:** 10.1101/2023.04.02.23288057

**Authors:** Debjyoti Talukdar, Luis Felipe de Deus, Nikhil Sehgal

**Author notes:** **Corresponding Author:** Dr. Debjyoti Talukdar, Department of Medical Research, Armenian Russian International University “Mkhitar Gosh”.

## Abstract

**Background:** Many research studies seek to improve vital sign monitoring to enhance the conditions under which doctors and caregivers track patients ‘ health. Non-invasive and contactless monitoring has emerged as an optimal solution for this problem, with telemedicine, self-monitoring, and wellbeing tools being the next generation of technology in the biomedical field. However, there is worldwide concern about the general purpose and bias towards a certain demographic group of these techniques. In particular, skin tone and the accuracy of monitoring dark skin tone groups have been key questions among researchers, with the lack of results and studies contributing to this uncertainty.

**Methods:** This paper proposes a benchmark for remote monitoring solution against a medical device across different skin tone people. Around 330 videos from 90 different patients were analyzed, and Heart Rate and Heart Rate Variability were compared across different subgroups. The Fitzpatrick scale (1-6) was used to classify participants into three skin tone groups: 1 and 2; 3 and 4; 5 and 6.

**Results:** The results showed that our proposed methodology was able to estimate heart rate with a mean absolute error of 3 bpm across all samples and subgroups. Moreover, for Heart Rate Variability (HRV) metrics, we achieved the following results, in terms of Mobility Assistive Equipment (MAE): HRV-IBI (Inter-Beat-Interval) of 10 ms; HRV-SDNN (Standard Deviation of Normal to Normal heartbeats) of 14 ms and HRV-RMSSD (Root Mean Square of Successive Differences between normal heartbeats) of 22 ms. No significant performance decrease was found for any skin tone group, and there was no error trend towards a certain group.

**Conclusions:** The study showed that our methodology meets acceptable agreement levels for the proposed metrics and is well-suited for users who want to understand their general health and wellness. Furthermore, the experiments showed that skin tone had no impact on the results, which remained within the same range across all groups.

## 1. Introduction

Non-invasive vital sign monitoring is an important aspect of healthcare as it allows doctors and caregivers to track a patient ‘s vital signs without the need for invasive procedures. Well-established methods for capturing physiological data include the use of the electrocardiogram (ECG), photoplethysmography (PPG), and bioelectrical impedance analysis (BIA), all of which require the use of contact sensors. These methods can measure several different physiological parameters, such as heart rate (HR), respiration rate (RR), oxygenation (SpO2), and blood pressure (BP) through their physiological signals [1–3].

Although these techniques have greatly increased the capabilities of not only doctors and physicians but also normal users, they still require contact with the patient through the use of sensors placed on the patient. However, researchers have introduced a technology that has shown promising results in this area: remote photoplethysmography (rPPG). This technology uses images from a regular camera combined with machine learning algorithms to estimate the same vital signs as contact-based methods.

The information acquired through rPPG reflects variations in blood volume in skin tissue, which are modulated by cardiac activity. The reflection of light is influenced by changes in blood volume and movement of the wall of blood vessels, and this phenomenon is visible through frame-to-frame changes in an RGB camera [4].

One key advantage of rPPG is that it is non-invasive and can be performed remotely, making it well-suited for monitoring patients in a variety of settings, including hospitals, homes, and the field. Additionally, rPPG is relatively low-cost, making it accessible to a wide range of healthcare providers.

However, there are several challenges to extracting an optimal rPPG signal. Distortions in the signal may be caused by low illumination, significant head movement, and device properties in low-end cameras such as frame rate and resolution. Moreover, pigmentation of the skin has been a major concern due to the light-based nature of the rPPG technique and the lack of studies involving participants with different skin tones.

To build an rPPG system, a four-step methodology is required, which can be summarized as frame-to-frame extraction, region of interest (ROI) detection, signal processing, and vital sign estimation. These steps are divided into blocks, all of which are based on well-established, state-of-the-art concepts such as face detection, landmark positioning, frequency spectrum analysis, and digital signal processing.

First, the video obtained from the camera is separated into several frames, with the number of frames per second denoted as the frame rate (FPS). One constraint of rPPG systems is the minimum FPS required to pick up fast changes in the cardiac cycle. As the heart rate increases, so does the required FPS, but this is usually not an issue for most existing smartphone cameras.

The next step is to find ROIs, which are usually extracted from the user ‘s face and theoretically compared to small sensors placed on the face. The position of the ROIs can vary among authors, but a common approach is to detect face regions in each video frame using face tracking algorithms such as the Viola-Jones method [5]. Once the ROIs are selected, pixel intensity components are extracted in the RGB color space. Furthermore, the RGB components are spatially averaged over all pixels in the ROI to yield a red, blue, and green component for each frame, forming the raw signals.

Additionally, signal processing is applied to the raw signal, which is also known as the “rPPG Core”. The rPPG Core has been the subject of various studies in the last decade, resulting in multiple methods that aim to extract a clean rPPG signal from the RGB components. There are many approaches in the literature to achieve this, such as those that rely on Blind Source Separation (BSS) methods [6,7]. These methods can retrieve information by de-mixing raw signals into different sources, including Principal Component Analysis (PCA)-based and Independent Component Analysis (ICA)-based techniques, which use different criteria to separate temporal RGB traces into uncorrelated or independent signal sources. Other authors have tried to improve the quality of the signal by changing the color space to a chrominance-based domain [8].

In a more recent study, Wang *et al*. [9] introduced a new alternative to process RGB components into rPPG signals, the “plane-orthogonal-to-skin” (POS) algorithm. In short, the POS method seeks to filter out intensity variations by projecting RGB components onto a plane orthogonal to a normalized skin tone vector. A 2-D signal referencing the projections is obtained and then combined into a 1-D signal, which is one of the input signal dimensions that is weighted by an alpha parameter. The alpha parameter is the quotient of the standard deviations of each signal.

Despite the successful results achieved by all of the aforementioned authors, none of them have evaluated or investigated the impact of skin tone on the results.

In this paper, we propose to evaluate rPPG Software Development Kit (SDK) version 3.0 against medical devices for participants with different skin tones. This SDK can be integrated into Android and iOS mobile apps as well as web applications and provides estimations of physiological assessments for a variety of vital signs, with Heart Rate and Heart Rate Variability (HRV) selected as the main features for this benchmark. The initial hypothesis of our methodology is to achieve a Heart Rate error within 3 bpm, HRV-IBI within 50 ms, and HRV-SDNN within 15 ms. Additionally, this paper seeks to address the assumption that rPPG technology suffers from bias when used on people with dark skin tones.

The remainder of this paper is structured as follows: Section 2 provides complete description of the methodology used in this study. Section 3 presents the obtained results for Heart Rate and Heart Rate Variability against medical devices for participants with different skin tones. Moreover, section 4 includes a discussion of the presented results and analyses that seek to evaluate the proposed methods. Lastly, section 5 revisits the most important points of this work and presents a conclusion.

## 2. Materials and Methods

In this chapter, we present an overview of rPPG technology, covering its most important aspects and blocks. Additionally, the methodology used in this study will be addressed. Fig. 1 shows the block diagram of the proposed study.

**Figure 1.**
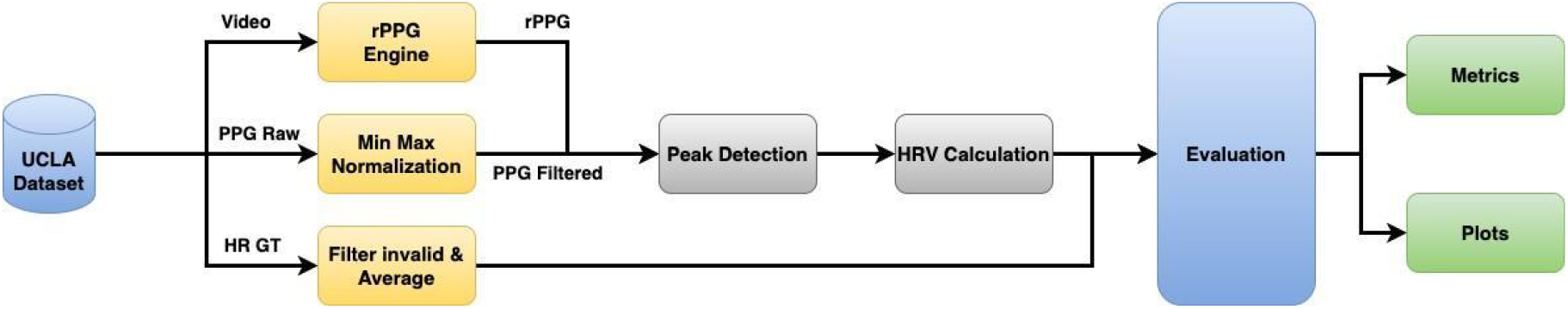
Block diagram of study methodology.

### 2.1. rPPG Processing

In this section, a detailed description of the rPPG core method will be provided. The aim of this method is to extract an optimal rPPG signal, which is as clean as possible and can contain the same physiological information as a PPG signal from a contact sensor. This signal reflects the cardiac cycle and body hemodynamics. The method is depicted in Fig.2.

**Figure 2.**
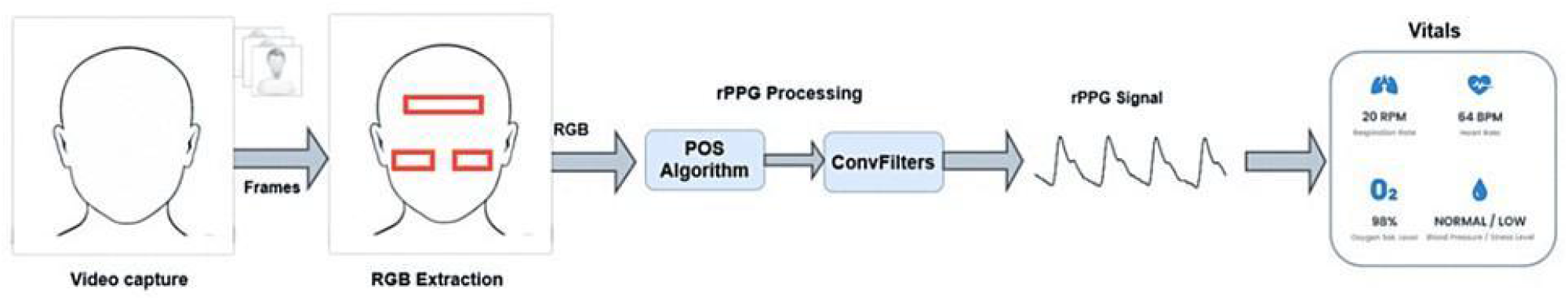
rPPG extraction method [11].

The RGB components were extracted from the ROIs using the landmark detection algorithm from the OpenCV library [10]. In a previous work [11], we proposed using three ROIs from the forehead, left cheek, and right cheek, which have shown to provide the best performance in internal experiments. Once the raw signal was collected, a version of the POS algorithm proposed by [9] is applied, and the resulting signal is further sent to a filtering stage based on convolutional filters. This stage aims to enhance the quality of the rPPG signal by denoising it as a sinusoidal wave.

#### 2.1.1. POS Algorithm

Originally proposed by Wang *et al*. [9], the POS algorithm seeks to mix RGB channels into a single-channel rPPG signal. According to the authors, the input RGB signal channels are mixed on the time interval *t* as following:

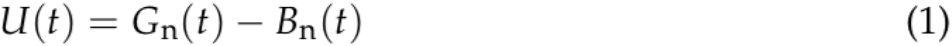

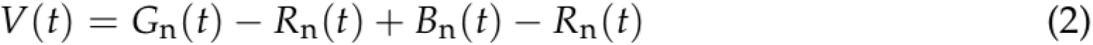

The subscript *n* stands for normalized, representing the instant color values divided by the mean value of the color channel.

The rPPG signal on this interval is constructed as denotes Eq.3:

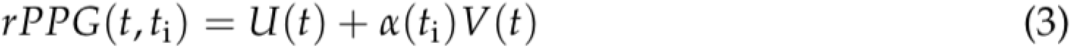

where *α* is the ratio between the standard deviation of U(*t*) and V(*t*) calculated on the interval.

#### 2.1.2. Convolutional Filter

The goal of this step is to enhance the signal quality by minimizing noise as much as possible, achieved by applying a convolutional filter (ConvFilter). The ConvFilter involves performing the convolution operation between the input single-channel signal, *s_orig*, which is extracted after the POS algorithm, and a template that represents a single heartbeat peak of the same signal. To construct the template, segments of *s_orig* signal around the detected peaks are averaged. Additionally, since *s_orig* signal could contain some noise, a band-pass filter with a bandwidth from 0.7 Hz to 7.0 Hz was also applied to facilitate peak detection.

The cleaner “s_heart” signal is obtained through convolution Eq.4 or the equivalent correlation Eq.5 with this template *t*[*k*]:

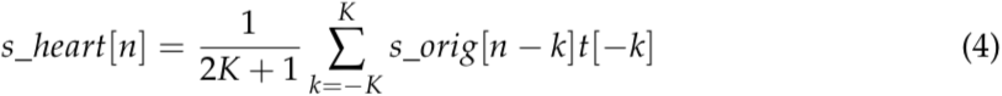

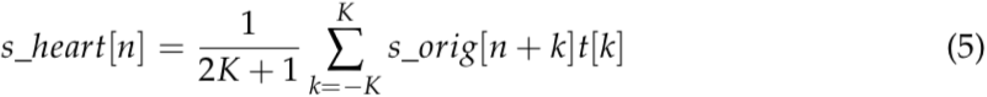

### 2.2. UCLA Dataset

In the past decade, rPPG technology has grown significantly with the advancement of computational power and the use of Machine Learning algorithms in various fields, including the biomedical field, where researchers have been improving non-invasive and contactless techniques for measuring physiological data. However, the increasing demand for benchmark datasets to evaluate these methods has also highlighted some concerns.

While many efforts have been made to collect rPPG datasets for more accurate physiological sensing, these datasets may have limitations, such as a small number of subject participants and biases toward certain demographic groups. Furthermore, few studies have explored the technology ‘s boundaries and limitations, such as its accuracy in darker skin tone populations, which remains largely unexplored due to the lack of proper datasets. For instance, Dasari et al. [12] proposed a dataset that only contains dark skin tones, but the actual videos are not shared, only the color space values of the skin region of interest.

Despite these challenges, Wang et al. [13] recently proposed the largest known rPPG dataset, which includes a variety of participants with different skin tones. The dataset comprises 98 subjects and 489 videos of various skin tones, ages, genders, ethnicities, and races. The skin tone of each subject was determined using the Fitzpatrick (FP) skin type scale [14], which ranges from 1 to 6. For each subject, five videos of approximately 1 minute were recorded at 30 frames per second (about 1800 frames), resulting in uncompressed videos with a total size of 2 gigabytes. All videos in the dataset are synchronized with ground truth heart rate and PPG signals extracted from a pulse oximeter placed on the subject ‘s finger.

### 2.3 Metrics

As mentioned earlier, the goal of this work is to evaluate the proposed method of remote health screening using rPPG signals extracted from video files. To assess the performance, heart rate and HRV features were used. Heart rate is a well-established parameter that is familiar to most people and ranges from low to high. In contrast, HRV may sound unfamiliar to many, but it can provide important insights into a person ‘s health. HRV-SDNN was used to assess health status, while HRV-LF and HRV-HF were correlated to the autonomic nervous system through the sympathetic and parasympathetic branches [15,16].

As heart rate was analyzed, the GT value provided in the dataset was compared to the estimated heart rate. For the HRV assessment, the comparison was made with the HRV features calculated using the PPG GT signal. Thus, physiological features were taken from both contact and remote signals and compared.

The RR Interval, also known as pulse-to-pulse interval, was the main tool used to calculate those features. It is the time difference between two peaks in milliseconds (ms) (Eq.6). Furthermore, the following features were used: Inter-Beat-Interval (IBI) (Eq.7), Root Mean Square of Successive Differences between normal heartbeats (RMSSD) (Eq.8), and the Standard Deviation of Normal to Normal heartbeats (SDNN) (Eq.9), all of which are in the time domain. In the frequency domain, the power from the low-frequency band (LF) [0.04; 0.15]Hz and high-frequency band (HF) [0.15; 0.4]Hz was measured. All comparisons were conducted in terms of Mean Absolute Error (MAE), Mean Error (ME), and 2-D plots.

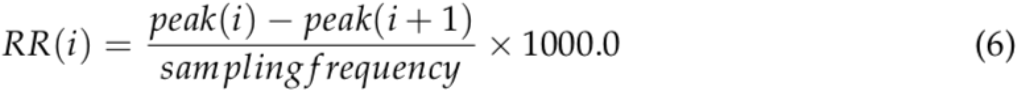

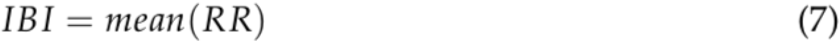

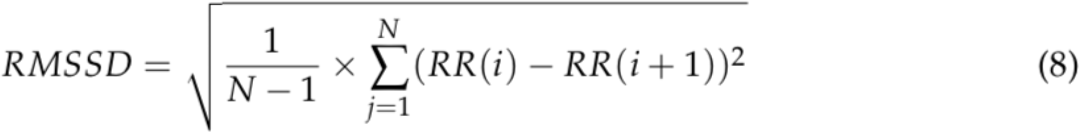

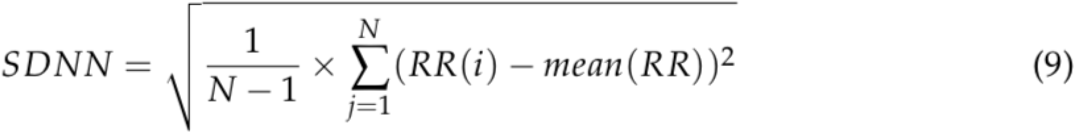

## 3. Results

In this study, video files from UCLA dataset [13] were used to extract physiological in- formation from individuals. Each video is processed using the proposed method described in section 2.1, which returns the rPPG signal. Furthermore, the information obtained through the rPPG signal is compared with the GT heart rate, as well as HRV features extracted from the contact PPG signal. The goal of this study is to not only benchmark the overall results but also assess the impact within certain groups based on skin tone and gender.

Firstly, after extracting the rPPG signals, it was necessary to remove some samples. However, the criteria differed for HR and HRV due to their different sources. Since HR was obtained directly from the sensor and HRV was calculated from the PPG signal, some samples might be useful for one benchmark but not for the other. The following issues were identified:

Some samples had no GT HR and were characterized by constant values of 255 and 129, which caused their exclusion from the dataset.

In addition, some samples were manually checked and three cases were observed: poor PPG GT quality, PPG GT discontinuity (probably caused by interference or sensor displacement), and increasing heart rhythm. Samples from the first two cases were removed from the HRV benchmark but not from HR, as long as the GT HR presented reasonable values (HR < 200 bpm).

Lastly, some samples presented an irregular heart rhythm, with an increase of up to 10 bpm at some point during the experiment. This might be caused by several factors such as not being at rest when taking the measurement, participants speaking or moving during the reading, or deliberately increasing heart rate. Additionally, a less likely cause could be a correlation with cardiac diseases. These samples were not removed from the dataset, as this work understands that the technology should be able to pick up these changes as well. However, this represents a bigger challenge.

Additionally, samples that could not have their HR estimated were also removed, as this work understands that there is no point in outputting a value if there is no certain confidence.

As a result, the dataset for HR benchmarking contained 339 samples from 90 unique participants, while the HRV dataset contained 332 and 94 samples, respectively. Fig. 3 shows a histogram of GT HR for all remaining samples.

**Figure 3.**
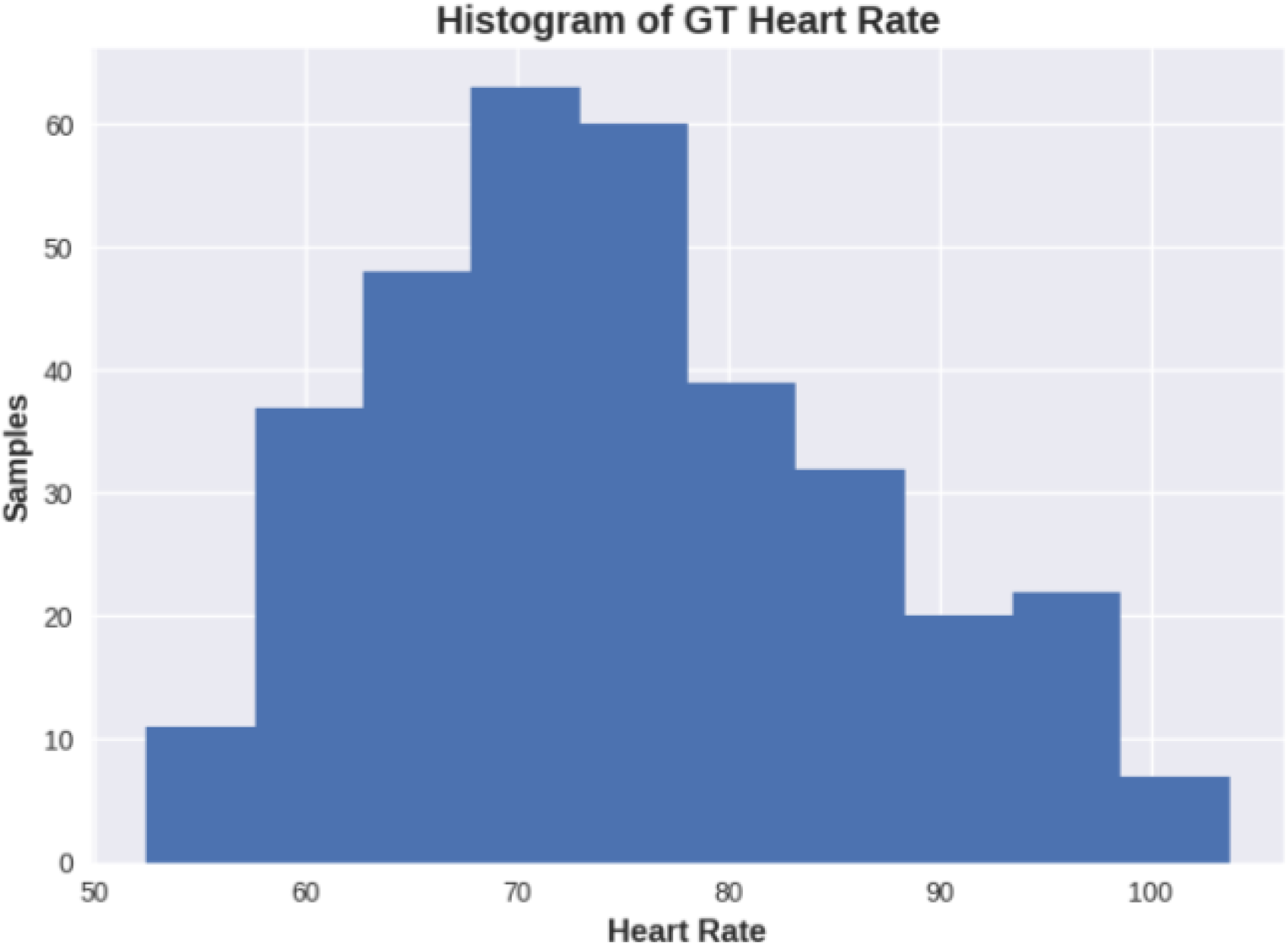
Ground Truth Heart Rate distribution.

For this study, subgroups were created to enhance the understanding of the results. Firstly, we evaluated the results from a signal quality perspective in terms of Signal to Noise Ratio (SNR), measured in decibels (dB). Three groups were used: SNR > 5 dB, which is classified as the minimum required, SNR > 8 dB as optimal signals, and SNR > 10 dB considered perfect signal quality. Regarding skin tone groups, based on the Fitzpatrick skin type [14], three other groups were considered: light skin tones related to values 1 and 2 of the scale, medium skin tones, consisting of skin tones with values 3 and 4 of the scale, and dark skin tones, consisting of skin tones 5 and 6 of the scale. Additionally, this study chose to split gender into two groups: male and female. For the skin tone and gender groups, no exclusion was performed based on minimum SNR.

Tab.1 shows the results for Heart Rate estimation for all samples as well as each one of the subgroups in terms of mean absolute error and mean error.

**Table 1:** Heart Rate evaluation across different subgroups

| Group | N. Samples | N. Subjects | Metrics (bpm) |  |
| --- | --- | --- | --- | --- |
|  |  |  | MAE | ME |
| All samples | 339 | 90 | 3 | 2 |
| SNR > 5 | 326 | 88 | 3.01 | 2 |
| SNR > 8 | 308 | 87 | 3.02 | 2.03 |
| SNR > 10 | 248 | 80 | 2.89 | 1.86 |
| Fitzpatrick [1,2] | 104 | 27 | 2.53 | 1.58 |
| Fitzpatrick [3,4] | 183 | 48 | 3.05 | 1.82 |
| Fitzpatrick [5,6] | 52 | 15 | 3.79 | 3.46 |
| Male | 116 | 29 | 3.24 | 2.01 |
| Female | 223 | 61 | 2.88 | 1.99 |

Additionally, Fig.4 presents two scatter plots: Fig.4a with all available samples (after filtering), and Fig.4b, which only includes samples with perfect signal quality (SNR > 10 dB). The figures depict the best fit line (black) and the perfect line (red).

**Figure 4.**
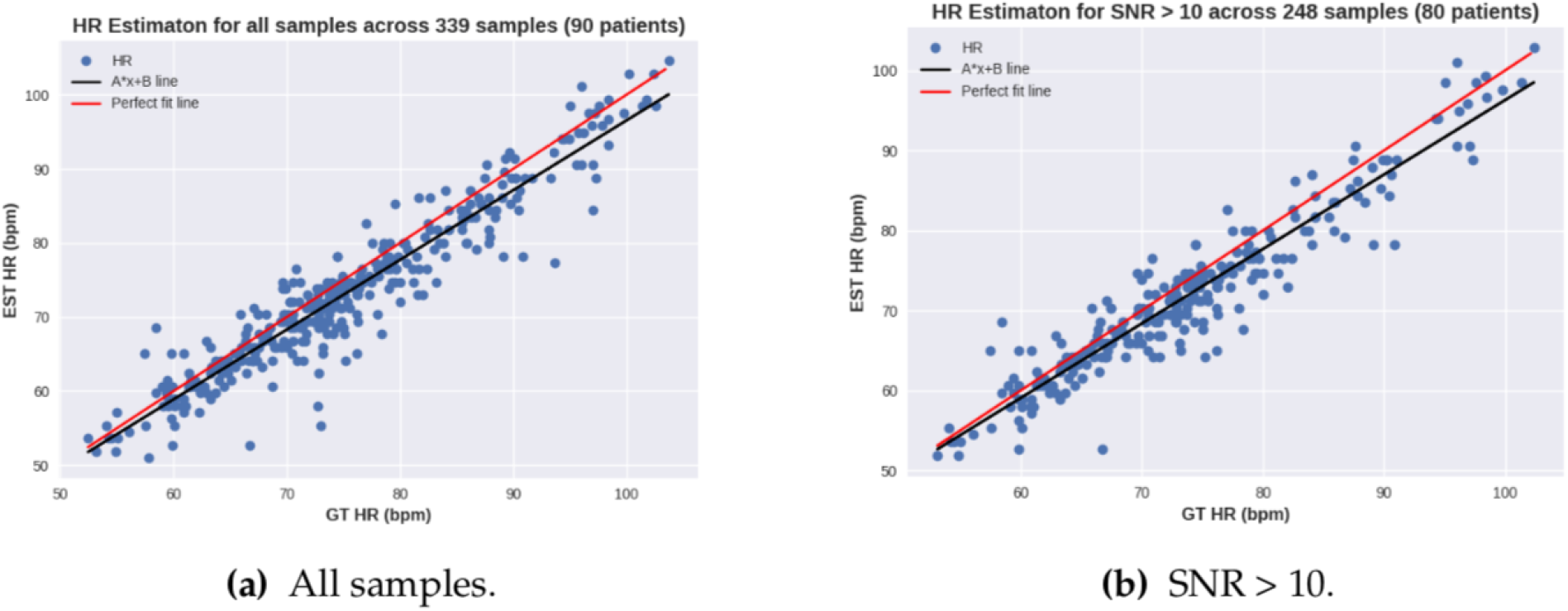
Scatter plot of HR across all samples and optimal signal quality.

Additionally, Fig.5 shows a similar plot for different skin tone groups. Fig.5a depicts people with light skin tones, defined by Fitzpatrick scale values 1 and 2, while Fig.5b displays the impact on medium skin tones for Fitzpatrick values 3 and 4. Finally, Fig.5c presents the results for dark skin tones, categorized as 5 and 6 on the Fitzpatrick scale.

**Figure 5.**
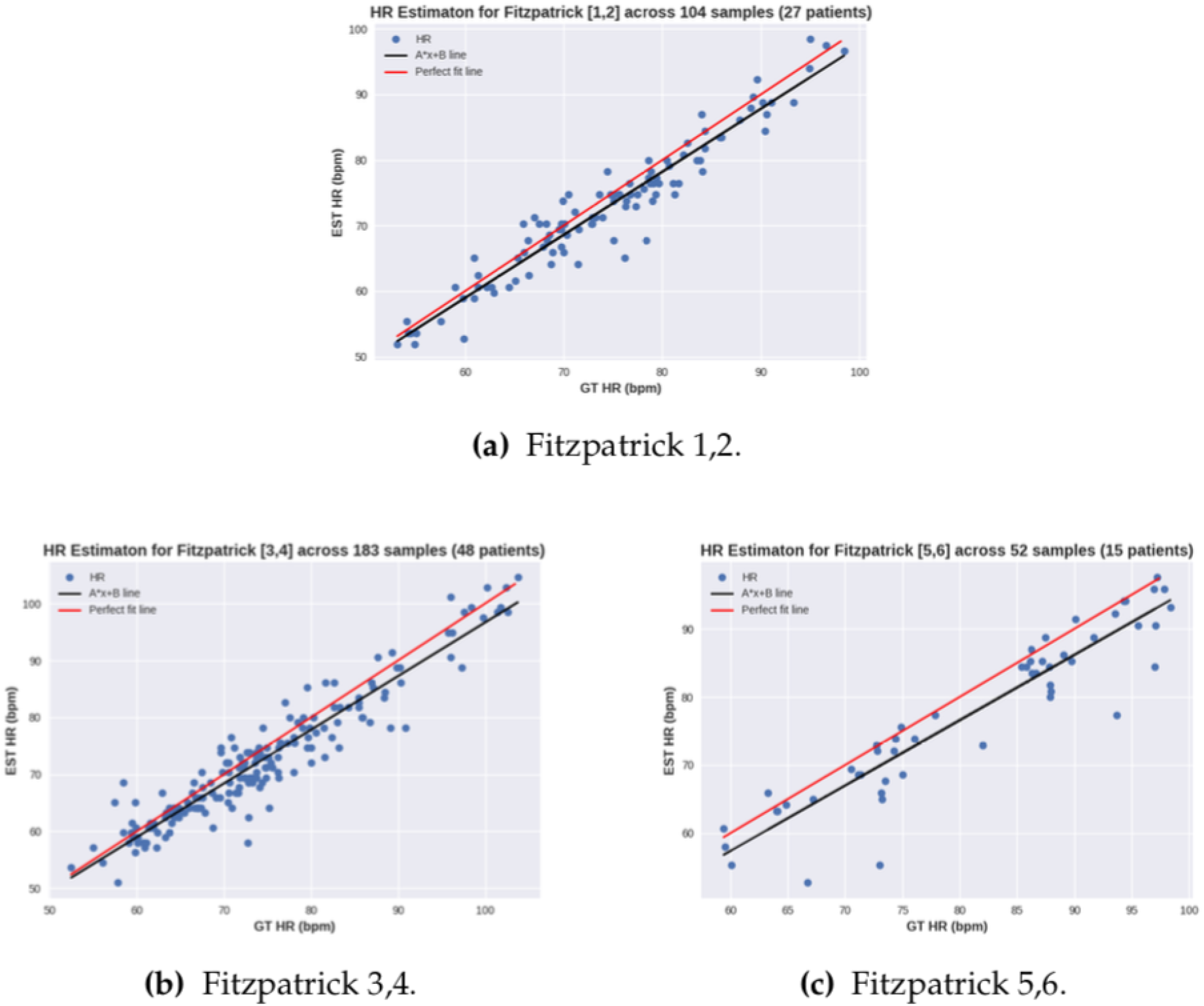
Scatter plot of HR for various skin tones according Fitzpatrick scale. a) skin tones 1 and 2; b) skin tones 3 and 4; c) skin tones 5,6.

Despite scatter plots being a fair representation of the distribution of errors, Bland Altman plots bring the results in terms of the mean value versus the difference between the ground truth (GT) and estimation. Fig.6 shows the Bland Altman plot for heart rate estimation across samples with SNR > 10 dB, where the skin tones were highlighted for each group.

**Figure 6.**
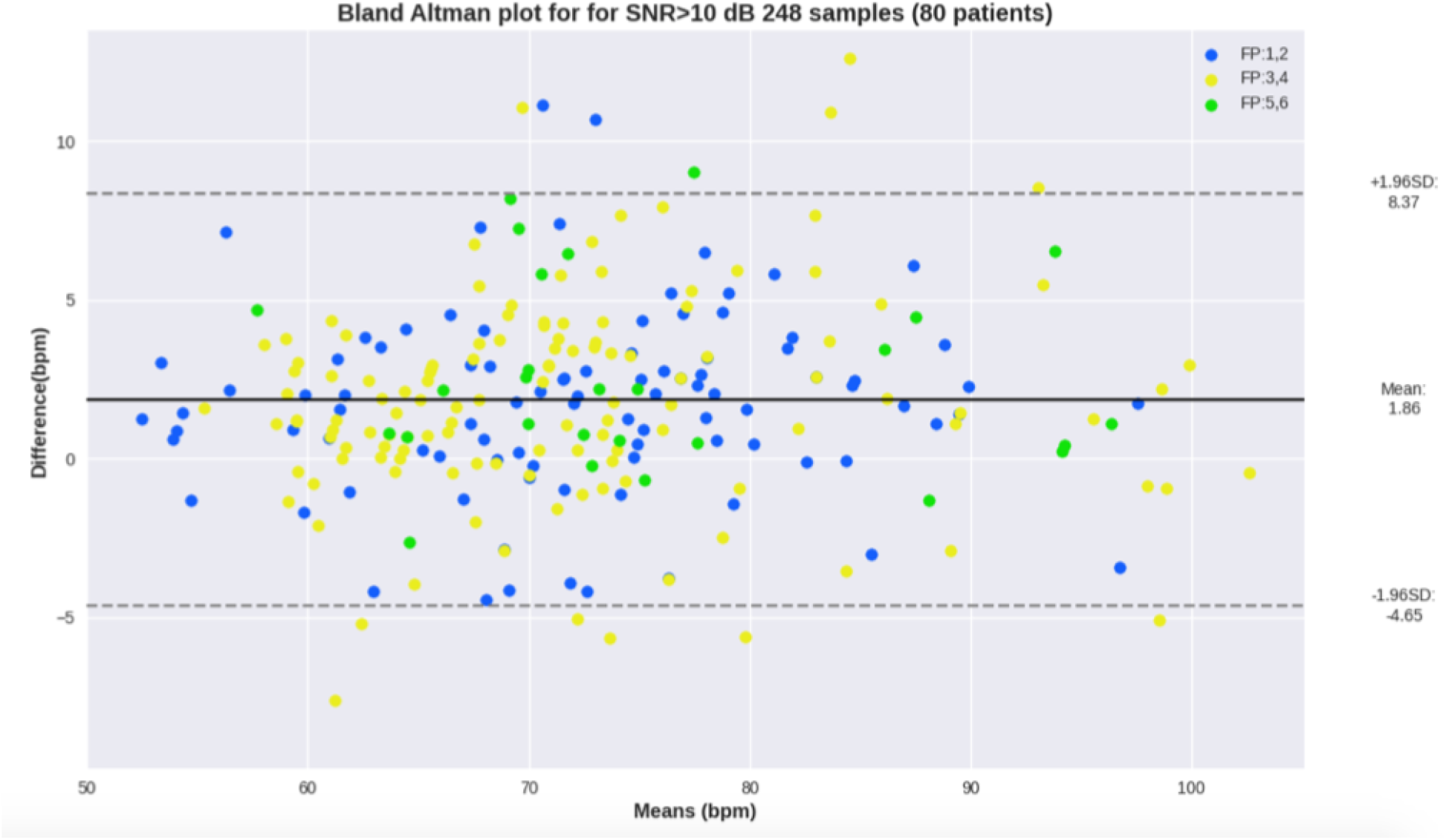
Bland Altman plot for Heart Rate estimation across samples with SNR > 10 dB.

Although heart rate is a reliable feature to benchmark physiological data extraction, heart rate variability (HRV) can provide a more in-depth analysis of someone ‘s current health status. As stated previously, both time and frequency domain HRV features can reveal important health insights. Similar to the heart rate analysis, Tab.2 and Tab.3 show an overview of the results, respectively, for the time and frequency domains, for all samples as well as for each subgroup.

**Table 2:** Heart Rate Variability time domain evaluation across different subgroups

|  |  |  | Metrics (ms) |  |  |  |  |  |
| --- | --- | --- | --- | --- | --- | --- | --- | --- |
|  |  |  | MAE |  |  | ME |  |  |
| Group | Samples | Subjects | IBI | SDNN | RMSSD | IBI | SDNN | RMSSD |
| All samples | 332 | 94 | 9.49 | 14.35 | 22.49 | -5.29 | 1.6 | 1.38 |
| SNR > 5 | 298 | 89 | 7.87 | 13.57 | 21.96 | -5.06 | 1.87 | 2.4 |
| SNR > 8 | 265 | 84 | 7.14 | 12.88 | 20.79 | -4.92 | 2.14 | 3.14 |
| SNR > 10 | 207 | 75 | 6.94 | 12.59 | 21.19 | -5.03 | 1.82 | 3.69 |
| Fitzpatrick [1,2] | 96 | 27 | 8.91 | 14.53 | 24.14 | -7.72 | 2.96 | 4.39 |
| Fitzpatrick [3,4] | 173 | 48 | 8.6 | 14.13 | 22.37 | -4.25 | 1.16 | 1.15 |
| Fitzpatrick [5,6] | 63 | 19 | 12.82 | 14.69 | 20.31 | -4.42 | 0.71 | -2.58 |
| Male | 111 | 30 | 9.62 | 15.54 | 21.29 | -4.67 | 3.88 | 1.23 |
| Female | 218 | 63 | 9.43 | 13.89 | 23.34 | -5.55 | 0.44 | 1.43 |

**Table 3:** Heart Rate Variability frequency domain evaluation across different subgroups

|  |  |  | Metrics (s<sup>2</sup>/Hz) |  |  |  |
| --- | --- | --- | --- | --- | --- | --- |
|  |  |  | MAE |  | ME |  |
| Group | Samples | Subjects | LF | HF | LF | HF |
| All samples | 329 | 94 | 16.2 | 34.09 | -5.26 | 9.67 |
| SNR > 5 | 295 | 89 | 15.97 | 33.8 | -5.57 | 8.78 |
| SNR > 8 | 262 | 84 | 15.16 | 32.71 | -5.78 | 9.28 |
| SNR > 10 | 205 | 75 | 15.66 | 33.49 | -6.48 | 9.32 |
| Fitzpatrick [1,2] | 96 | 27 | 17.34 | 28.02 | -4.68 | 8.53 |
| Fitzpatrick [3,4] | 170 | 48 | 14.62 | 35.93 | -6.2 | 7.39 |
| Fitzpatrick [5,6] | 63 | 19 | 18.69 | 38.38 | -3.6 | 17.56 |
| Male | 108 | 30 | 15.84 | 31.76 | -1.9 | 1.39 |
| Female | 218 | 63 | 16.37 | 35.43 | -7.19 | 13.69 |

This work proposes two main analyses of heart rate variability (HRV) for a more indepth assessment of participants ‘ health status. Firstly, an analysis of the inter-beat-interval (IBI) across all samples is presented in Fig.7, which highlights the mean IBI and thus the cardiac rhythm.

**Figure 7.**
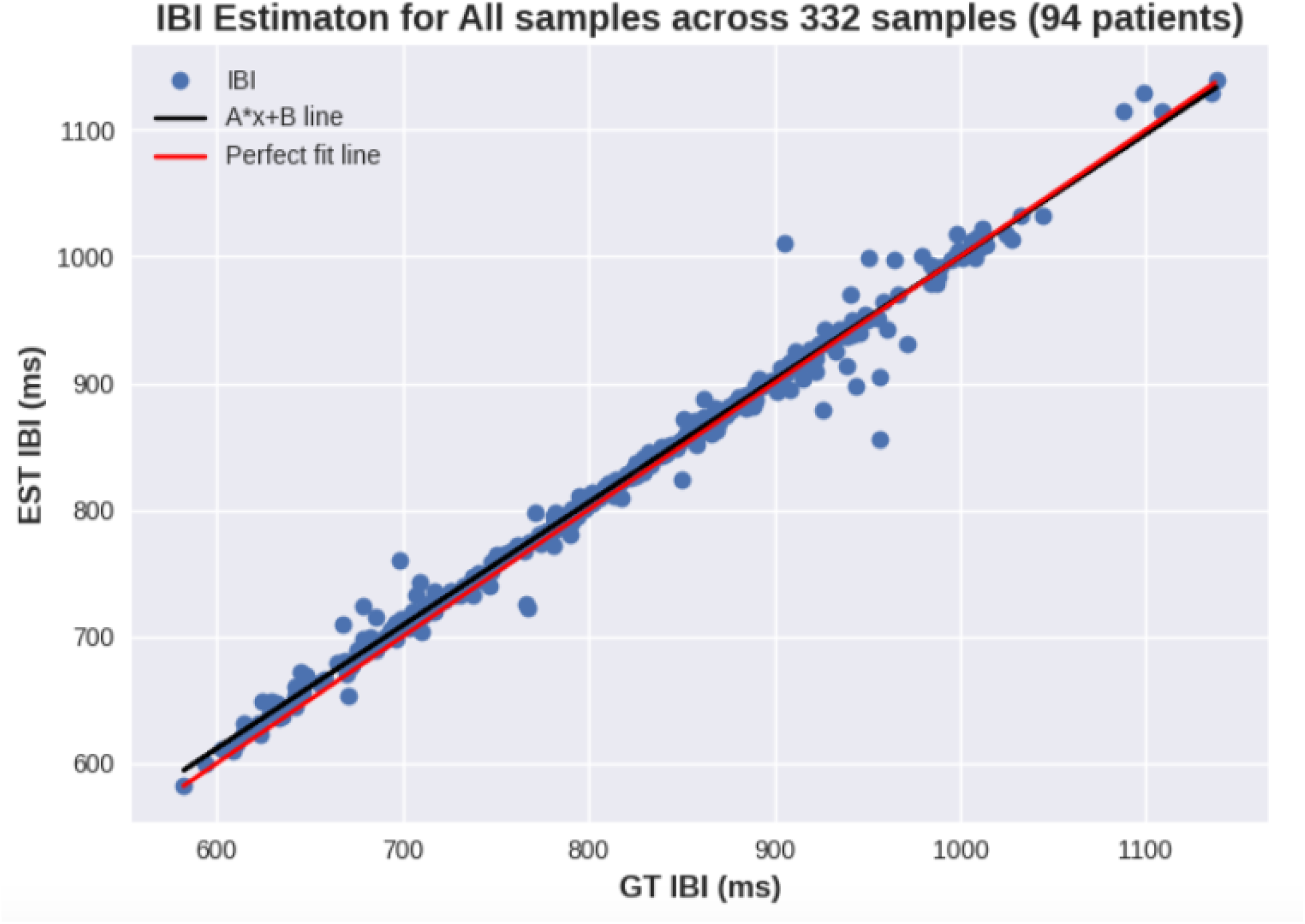
Scatter plot of Inter-Beat-Intervals across all samples.

Secondly, the study has chosen SDNN as the main HRV metric for in-depth analysis, as SDNN has been used as a fitness and health score. Fig.8 shows a scatter plot of SDNN across all samples (Fig.8a) as well as for samples with perfect signal quality (Fig.8b). For these plots, the skin tone groups were highlighted in different colors to identify the impact of skin tone on the presented errors. Similarly, Fig.9 shows the scatter plots for each skin tone group separately. Lastly, the Bland Altman plot for SDNN across samples with SNR > 10dB is presented in Fig.10. As with the previous plots, skin tone groups based on the Fitzpatrick scale are highlighted in different colors to facilitate the identification of samples with higher errors.

**Figure 8.**
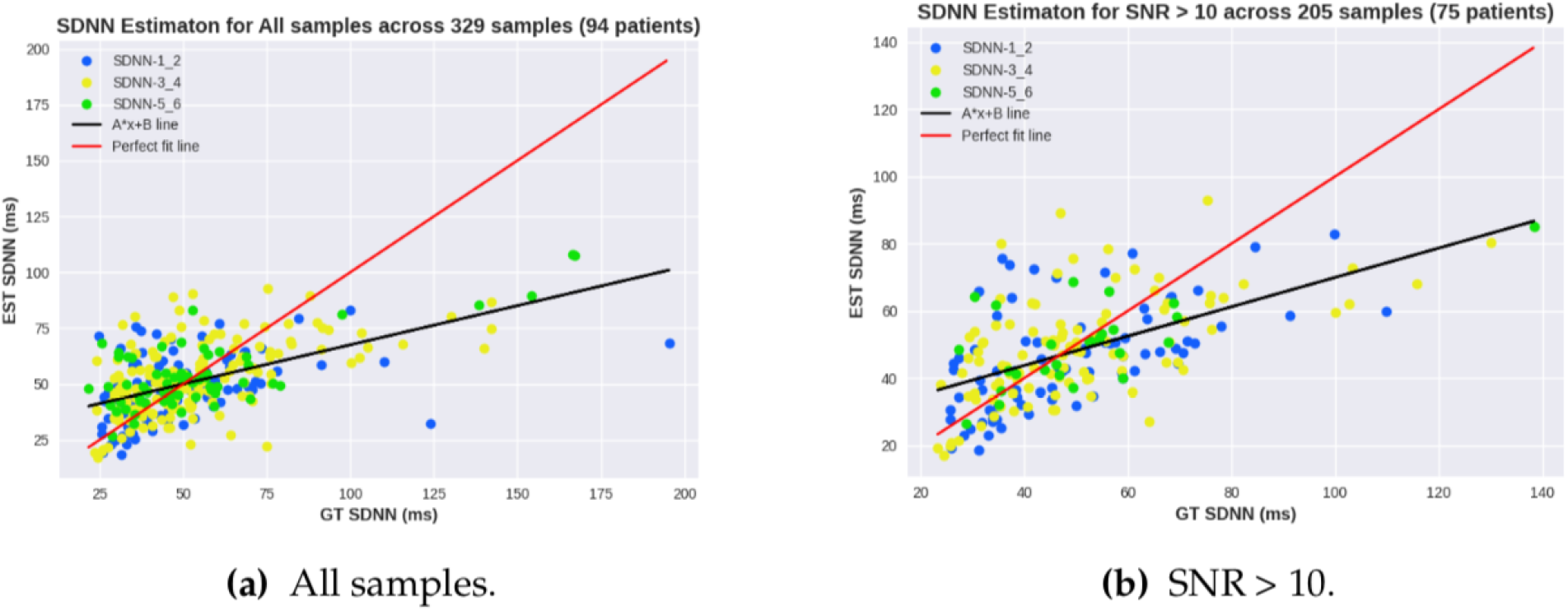
Scatter plot of SDNN across all samples and optimal signal quality

**Figure 9.**
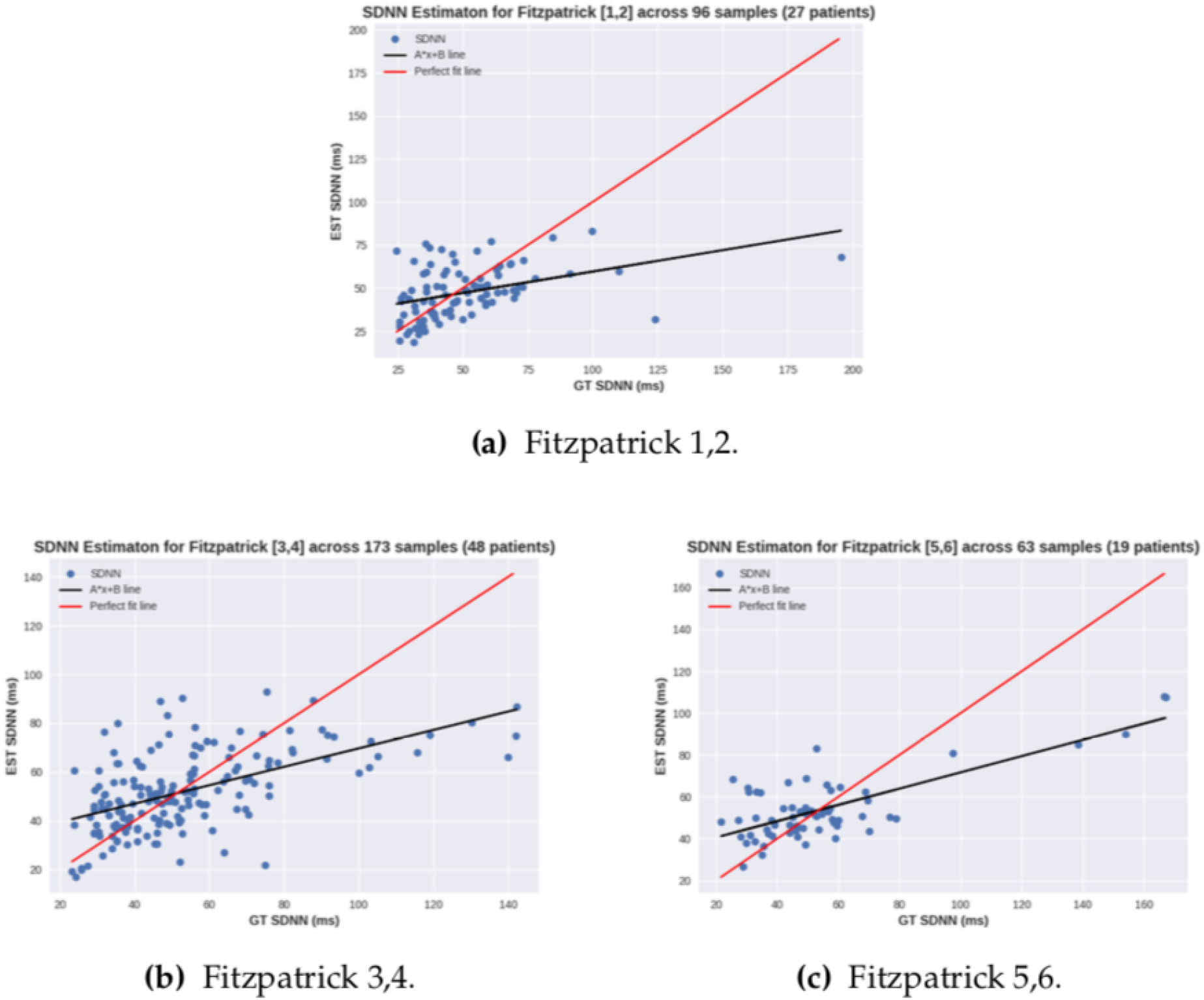
Scatter plot of SDNN for various skin tones according to Fitzpatrick scale. a) skin tones 1 and 2; b) skin tones 3 and 4; c) skin tones 5,6.

**Figure 10.**
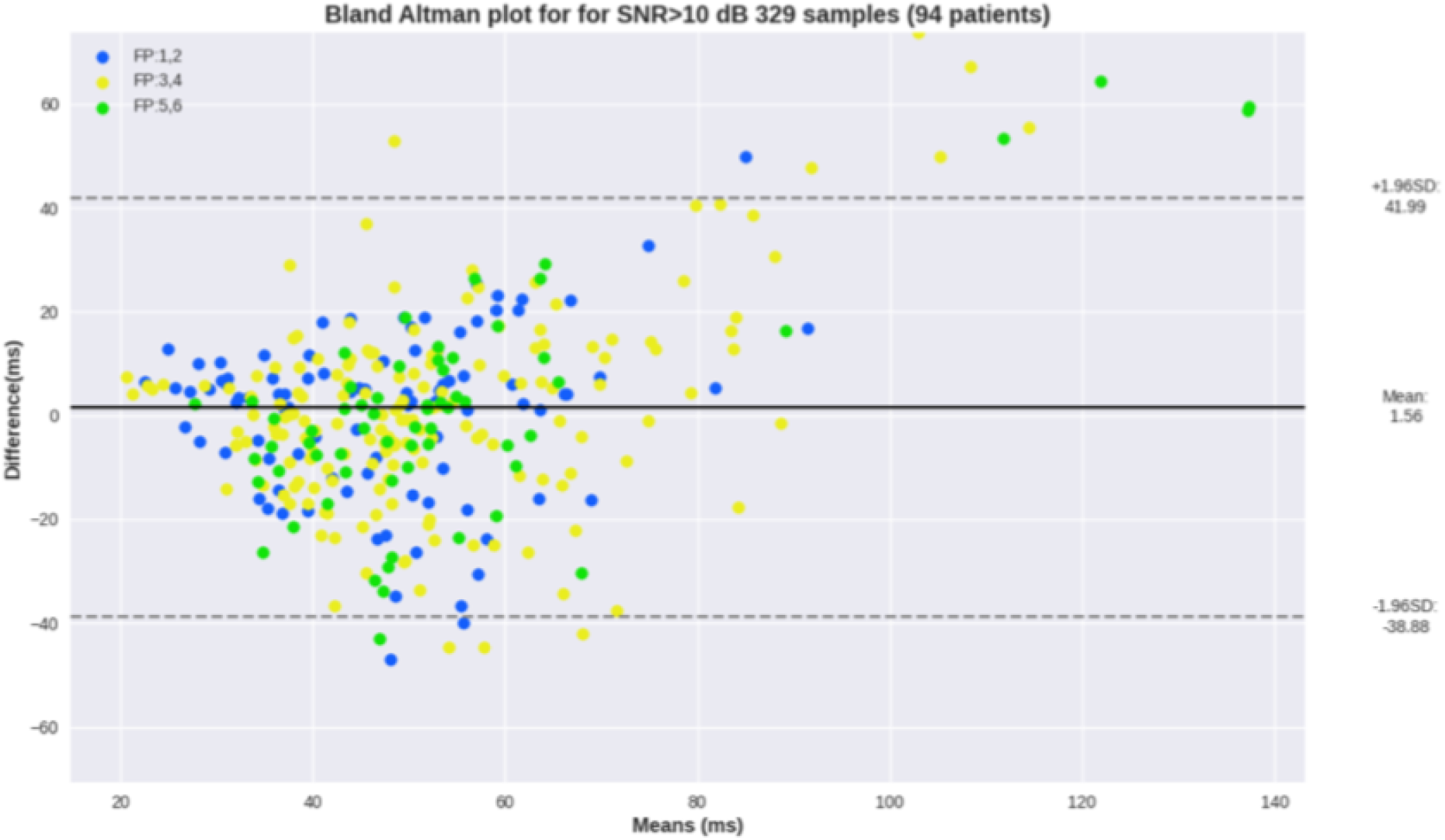
Bland Altman plot for SDNN estimation across samples with SNR > 10 dB.

## 4. Discussion

In this work, video files of 98 participants were extracted from the UCLA dataset, resulting in a total of 498 videos, each lasting 1 minute and captured at a frequency of 30 Hz. For each video, contact PPG and heart rate were synchronously measured using a pulse oximeter, and the skin tone was classified using the Fitzpatrick scale. After verifying the integrity of the data and removing erroneous samples, around 330 samples from 90 patients were retained.

The benchmark was conducted in terms of heart rate and heart rate variability, which are well-established metrics used to assess an individual ‘s health status. This work proposed dividing the samples into subgroups to enhance the analysis perspective. First, in terms of signal quality, three groups were created for minimum, optimal, and perfect conditions. Additionally, the samples were divided into three skin tone groups based on the Fitzpatrick scale, as well as two gender groups.

The aim of the first experiment was to evaluate the accuracy of the algorithm for heart rate estimation. Tab.1 shows that across all samples, the MAE error was 3 bpm, which meets the initial hypothesis. Moreover, the performance was stable across samples with SNR > 5 and 8 dB, as can be seen in Fig.4. This indicates that even with lower signal quality, the algorithm can still estimate HR accurately. When taking into account signals with perfect quality, there is a slight improvement to 2.89 bpm. It is worth noting that more than half of the samples had perfect signals, as well as 80 out of 90 patients. Despite the fact that the dataset was collected in a good setting, signal quality is also a direct result of power on calibration of light changes and movement compensation to enhance the extracted signal.

With regard to skin tone groups, most of the participants (48) were in the medium skin tone group (FP 3,4), which achieved an MAE of 3.05 bpm. The dark skin tone group had a slightly higher MAE of 3.79 bpm, but it was less than 1 bpm from the original mark. Through the comparison between Fig.5c and Fig.5b, it is possible to see the difference in the relationship between the 1:1 line and the best fit line, where in the former, the greatest differences remain when the estimated value was smaller than the actual value.

Despite the initial results showing a slight decrease in performance, this may be due to other factors, such as signal quality and an increase in heart rate throughout the reading. The Bland-Altman plot presented in Fig.6 corroborates this hypothesis, as it shows all readings with SNR 10+ and no correlation can be seen between skin tone group and errors. The blue dots represent light skin tones, yellow for medium, and green for dark skin tones. Additionally, the plot shows a mean error of 1.86 bpm across all the readings with SNR > 10 dB.

Moreover, out of the 98 participants, 61 were women and 29 were men. There was no major discrepancy in the results between the two gender groups, indicating no bias towards gender.

Regarding the HRV experiments, this work extracted results from five different features, three of which are from the time domain (IBI, SDNN, and RMSSD), and two are from the frequency domain (LF and HF). In the overview results presented in Tab.2, it is possible to see that IBI displays the lowest error range within 10 ms, which is a significant result and far exceeds the initial hypothesis of 50 ms. If we consider that IBI ranges from 500 to 1500, 10 ms would reflect an average error of 1 bpm. Moreover, SDNN, as previously stated, has been used to determine the fitness score and shows an average error of 14 ms, which also meets the proposed value of 15 ms. Normal ranges of SDNN vary from 30 to 150.

Through the analysis of HRV, conclusions can be drawn about the previous experiment as well, since HRV can provide additional insight into the cardiac rhythm. While IBI displays an almost perfect correlation, as observed in Fig.7, SDNN shows some discrepancies, often related to high SDNN values as can be seen in Fig.8a, 9a, and 9c, where the ground truth (GT) SDNN was above 100 ms. These values are highly associated with the increasing heart rhythm previously discussed, which can be hidden in some metrics but clear in the standard deviation. While these values are not wrong in terms of metrics, they do not reflect the rhythm conditions, most likely because they were not at rest. Thus, these samples should be considered as outliers, and their exclusion would result in a decrease in MAE SDNN to around 9 ms.

Regarding subgroups, the overview results show that MAE was consistent across all Fitzpatrick groups (around 14 ms). The plots show sparse estimations, and once again, no correlation between the skin tone group and the errors can be seen. The Bland-Altman analysis in Fig.10 shows a mean SDNN error across samples with SNR 10+ of 1.56 ms, and most differences remain within +20 ms.

Additionally, an increase in the MAE of the male group to 15.54 ms can be observed. This phenomenon can be explained because most of the samples with an increasing heart rhythm were from men, which directly affects the SDNN and RMSSD metrics.

Regarding RMSSD, although most of the MAE results remain stable around 22 ms, it is noticeable that the lowest MAE is observed in the dark skin tone group, with 20.31 ms.

Finally, acceptable results were also obtained in the frequency analysis. The normalized power of the low and high frequency bands was extracted from contact and remote signals for comparison. As shown in Tab.3, the MAE for the low frequency band is within 16 *s*2/Hz, while for the high frequency band it is within 38 *s*2/Hz. The normal ranges for LF and HF are respectively 0-70 *s*2/Hz and 10-175 *s*2/Hz. Through the LF and HF features, correlations with the Autonomic Nervous System can be inferred.

## 5. Conclusions

In this paper, a benchmark of our methodology using the largest known rPPG dataset was proposed. Approximately 340 video files from 90 patients were analyzed, and from these, rPPG signals, heart rate, and heart rate variability were extracted and compared to ground truth information obtained from a pulse oximeter. The aim of this study was to address the lack of results of rPPG technology in a population of individuals with different skin tones, as well as to mitigate concerns about the accuracy of the method in people with dark skin tones.

Heart rate and heart rate variability were chosen as the main features to evaluate similarity due to their ease of extraction, well-known features, and ability to potentially reflect health and fitness insights about the user.

The analysis was carried out within subgroups based on skin tone, with three groups created based on the Fitzpatrick scale. The results have shown that the heart rate meets the initial hypothesis of a mean absolute error of 3 bpm. Within the skin tone subgroups, no significant performance glitches were observed. Moreover, the HRV results have shown an almost perfect correlation with IBI, with a mean absolute error within 10 ms (around 1 bpm). Similarly, SDNN has shown acceptable results within 14 ms, although some samples presented an increasing cardiac rhythm during the reading, which drove the HRV metrics towards deceptive values.

The study has shown that our methodology meets acceptable agreement levels for mean absolute error for HR, HRV-IBI, HRV-SDNN, HRV-RMSSD, HRV-LF, and HRV-HF. Furthermore, the experiments have shown that skin tone had no impact on the results, which all remained within the same range. Moreover, this work has demonstrated through its results that rPPG technology can be used and has no significant accuracy impact on dark-skinned people. This shows that the proposed methodology is acceptable for users who want to understand their general health and wellness.

## Data Availability

Researchers interested in access to the data may contact Nikhil Sehgal at, also see https://vastmindz.com/contact/. The authors will assist with any negotiable data use agreements and gain access to the data for any replication efforts following publication.

## Abbreviations

The following abbreviations are used in this manuscript:

AI: Artificial Intelligence
HR: Heart Rate
HRV: Heart Rate Variability
IBI: Inter-Beat-Interval
PPG: Photoplethysmography
RMSSD: Root Mean Square of Successive Differences between normal heartbeats SDNN Standard Deviation of Normal to Normal heartbeats

